# Maternal genetic risk for high body mass index and reduced infant mid-childhood educational attainment

**DOI:** 10.64898/2026.11.17.26368342

**Authors:** Tabitha Wishlade, Jasmine A Mack, Laurentya Olga, Ulla Sovio, Felix Day, Gordon Smith, Catherine E Aiken

**Author notes:** Corresponding author: Professor Catherine Aiken; Address: Department of Obstetrics and Gynaecology, University of Cambridge, Box 223, The Rosie Hospital and NIHR Cambridge Biomedical Research Centre, Cambridge CB2 0SW, United Kingdom.

## Abstract

**Background:** Maternal body mass index (BMI) ≥30kg/m^2^ has previously been associated with reduced infant neurocognitive development, including educational attainment. The mechanistic pathway underlying this association is unclear

**Objective:** We examined the relationship between observed maternal BMI, maternal genetic liability for BMI, and infant mid-childhood educational attainment. We aimed to limit residual confounding by environmental factors, for example socioeconomic status.

**Methods:** A BMI polygenic risk score (PRS_BMI_) was applied to eligible participants in the Pregnancy Outcome Prediction study (n=2437). Six educational outcomes through mid-childhood (age 5, age 6, and 4 domains at age 7) were regressed on (i) maternal pre-pregnancy BMI based on self-reported weight (ii) maternal PRS_BMI_ and (iii) residual BMI unexplained by PRS_BMI_. Participants were stratified into four risk groups by PRS_BMI_ risk and observed BMI. Relationships between each risk group and educational outcomes were examined. All association analyses were adjusted for confounders including school funding type, socioeconomic status, and serious childhood morbidity. Causality was evaluated using two-sample Mendelian randomisation (MR). False discovery rate (FDR) was controlled using the Benjamini-Hochsberg correction at alpha=0.05.

**Findings:** Inverse associations were present between all education outcomes and maternal pre-pregnancy BMI, maternal PRS_BMI_, and maternal residual BMI. Children whose mothers had both pre-pregnancy BMI >25kg/m^2^ and PRS_BMI_ z-score >0 were consistently less likely to meet educational standards compared to other risk groups. In two-sample MR an inverse relationship between maternal BMI and educational attainment was statistically significant for educational attainment aged 6, but not at ages 5 or 7 after FDR correction.

**Interpretation:** Increased maternal pre-pregnancy BMI shows both genetic and environmental associations with reduced childhood educational attainment, although the two pathways are unlikely to be discrete. Infants of women with co-occurring high observed BMI and high genetic liability to BMI may be at highest risk of poor educational attainment.

## Introduction

High maternal body mass index (BMI) (classified here as ≥30kg/m^2^)^(1)^ in pregnancy has previously been associated with reduced infant neurocognitive development^(2)^, ^(3)^,^(4)^, including mid-childhood educational attainment^(5)^. As over 16% of women globally enter pregnancy with BMI ≥30kg/m^2(6)^, even modest BMI-related effects on infant cognition could be impactful at a population level^(7)^. At an individual level, educational attainment at 7 years of age may predict mid-life socio-economic status, even when adjusted for childhood socio-economic status^(8)^, representing a potentially modifiable influence on long-term socio-economic mobility.

Biological mechanisms that have previously been suggested to underlie the association between maternal BMI and childhood neurocognition include fetal exposure to oxidative stress^(9)^, inflammation^(10)^, and elevated concentrations of leptin, glucose, and insulin^(11)^. These have been hypothesised to disrupt various aspects of fetal neurodevelopment^(4)^ *in utero* and thus may affect subsequent learning. Pregnancy conditions associated with high BMI, such as gestational diabetes and hypertensive disorders of pregnancy, may also be implicated^(12)^. However, establishing causality remains challenging: the association is highly susceptible to confounding by factors such as socioeconomic status, postnatal diet, maternal mental health and maternal cognition^(13)^. Methodological heterogeneity, for example in statistical modelling, neurocognitive assessment timing and method, and BMI assessment, may also explain inconsistent results between existing studies^(14)^,^(15)^.

We assessed whether the association between maternal BMI and childhood educational outcomes persisted when maternal genetic risk for BMI was taken into account. Our objectives were to examine associations between maternal polygenic risk score for BMI (PRS_BMI_) and educational attainment at ages 5 to 7 years, and to evaluate these associations for causality using two-sample Mendelian randomisation (MR).

## Methods and Materials

### Study population and data sources

The Pregnancy Outcome Prediction study (POPS) is a prospective UK cohort study recruiting from 2008 to 2012. The cohort comprises nulliparous women with viable singleton pregnancies recruited at their dating ultrasound scan (<14 weeks’ gestation) at the Rosie Hospital (Cambridge, UK)^(16)^,^(17)^.

Our analytic cohort included women from POPS with a livebirth, who were genotyped, had their pre-pregnancy BMI recorded, and had linked data to ≥1 standardised educational testing outcome. Educational outcomes were evaluated at 5, 6, and 7 years, using data from the National Pupil Database (NPD) curated by the UK Department for Education. Educational outcomes and assessment methods have been detailed elsewhere^(5)^.

### Outcomes

Our primary educational attainment outcome was Good Level of Development (GLD), assessed as standard for all children in publicly-funded UK schools (∼94% of the population resident in England^(18)^ at age 5. Secondary outcomes were educational attainment in phonics assessed at age 6, and domain-specific achievement in science, reading, writing and mathematics assessed at age 7.

GLD is a teacher-assessed composite measure of 12 early learning goals in (i) communication and language, (ii) physical development, (iii) personal, social, and emotional development, (iv) literacy, and (v) mathematics^(19)^. GLD is reported as a binary outcome corresponding to whether the expected standard was met.

At age 6, children undertook a 40-word phonics screening check one-on-one with a teacher. Children’s scores were compared to a national threshold to derive a binary outcome corresponding to whether the expected standard was met. Children who did not meet the expected standard aged 6 repeated the test aged 7^(5)^,^(20)^. Our analysis includes those children who retook the test, using whether the expected standard was ever achieved as the binary outcome measure.

At age 7, children were assessed as working towards, at or at greater depth than the expected standards in reading, writing, mathematics, and science by their class teacher^(21)^. From this, a binary outcome measure corresponding to whether the expected standard was met was generated for each domain^(5)^.

### Exposures and covariates

The primary exposure was maternal genetic risk for BMI. DNA from maternal blood was genotyped using the Illumina Infinium Global Screening Array Kit (GSA v3). The genotyping, quality control, kinship filtering, and imputation methods used are detailed elsewhere^(22)^. We applied a PRS_BMI_ including nearly 1.3 million variants^(23)^ from the PGS Catalog^(24)^ to eligible participants. To calculate the PRS_BMI_ we used the PGS Catalog calculator pipeline^(24)^, with continuous ancestry adjustment.

As our analytic cohort was ∼95% self-described white ethnicity (Table 1), for two-sample MR we used single nucleotide polymorphisms (SNPs) from publicly available summary data from the IEU OpenGWAS database from a meta-analysis of genome-wide association study (GWAS) BMI data in nearly 700 000 individuals of European ancestry^(25)^.

**Table 1:**
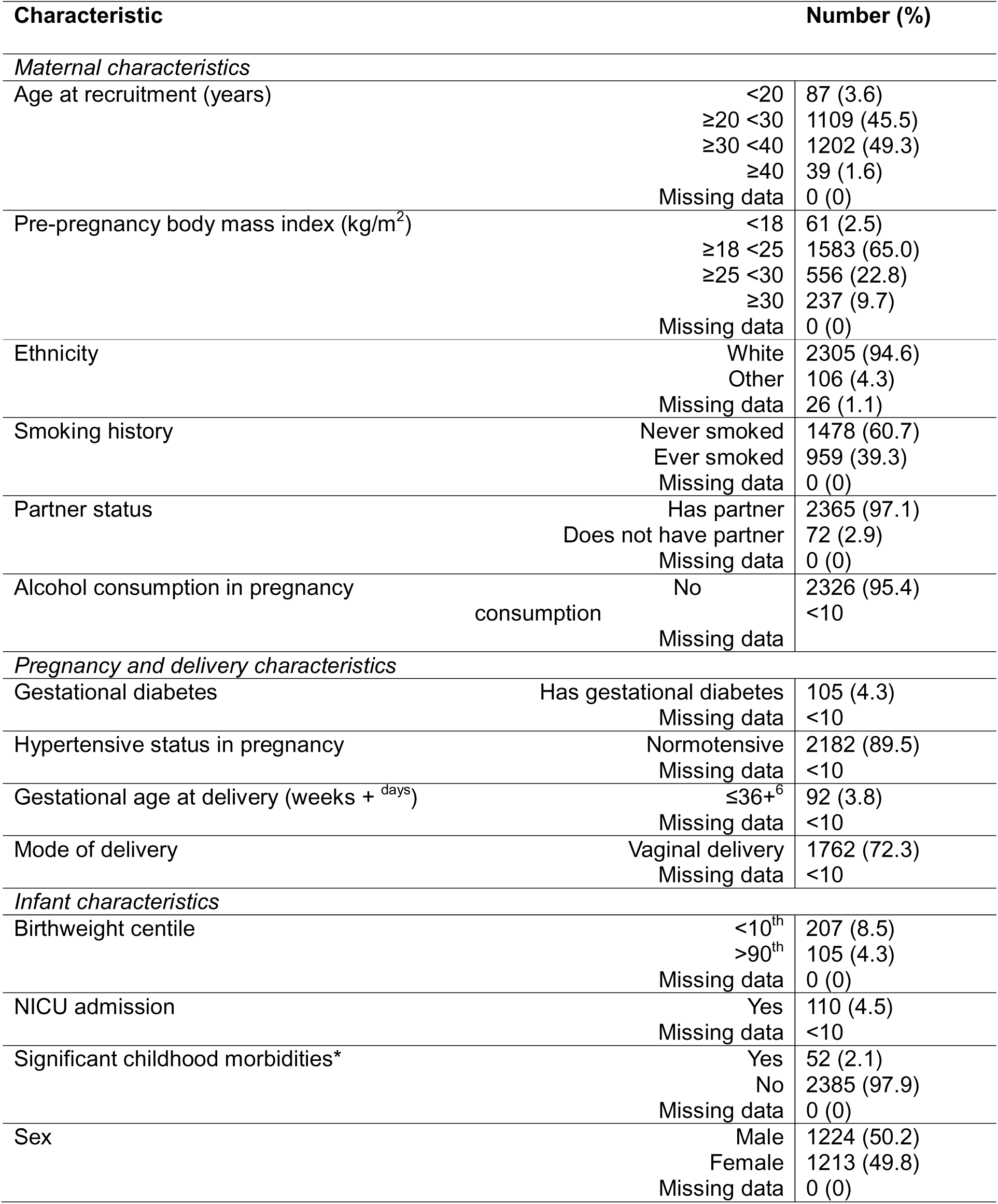
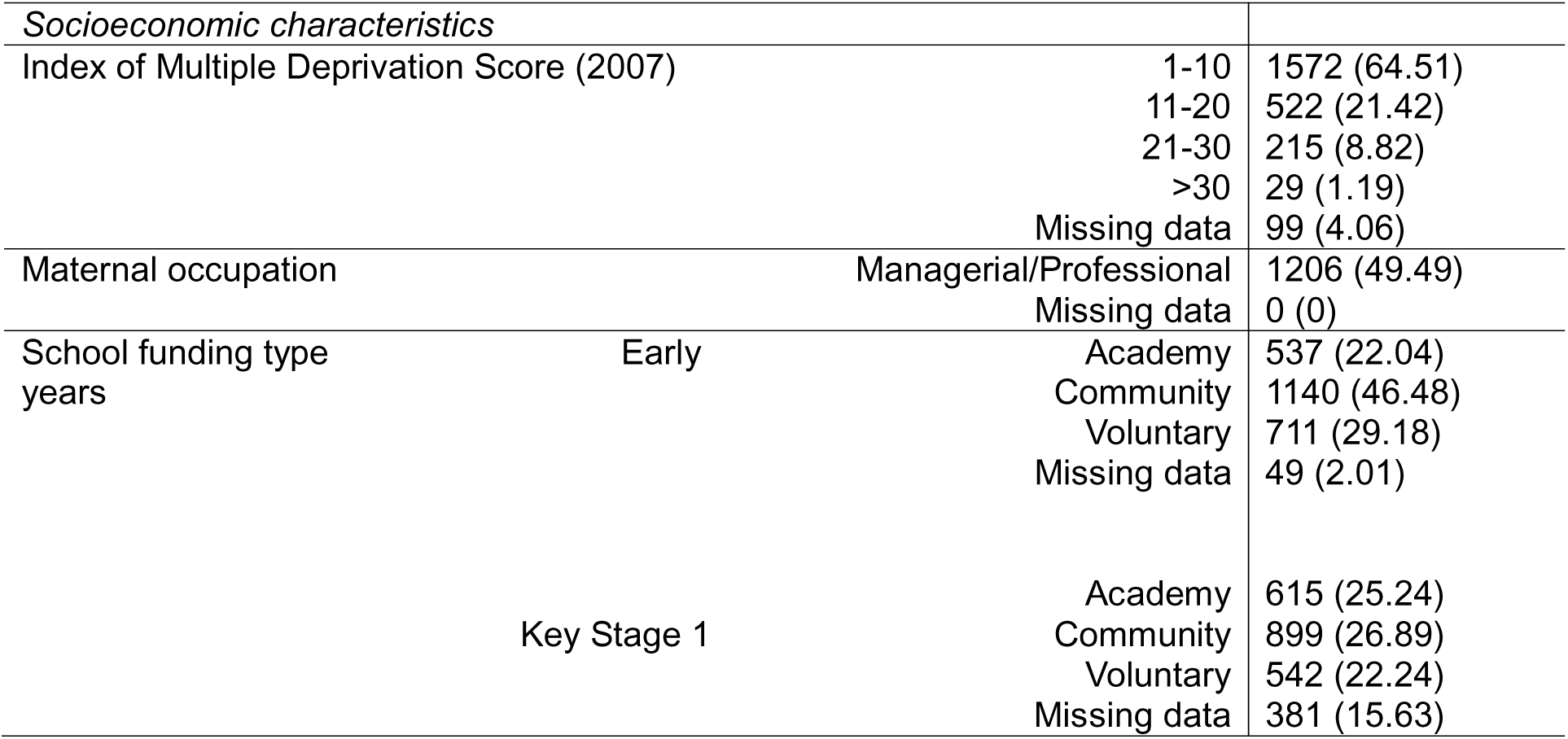
Demographics of analytic cohort. N = 2437. NICU = neonatal intensive care unit. Any numbers <10 are supressed to comply with the Office of National Statistics’ data presentation policy.

| Characteristic |  | Number (%) |
|---|---|---|
| *Maternal characteristics* |  |  |
| Age at recruitment (years) | <20 | 87 (3.6) |
|  | ≥20 <30 | 1109 (45.5) |
|  | ≥30 <40 | 1202 (49.3) |
|  | ≥40 | 39 (1.6) |
|  | Missing data | 0 (0) |
| Pre-pregnancy body mass index (kg/m<sup>2</sup>) | <18 | 61 (2.5) |
|  | ≥18 <25 | 1583 (65.0) |
|  | ≥25 <30 | 556 (22.8) |
|  | ≥30 | 237 (9.7) |
|  | Missing data | 0 (0) |
| Ethnicity | White | 2305 (94.6) |
|  | Other | 106 (4.3) |
|  | Missing data | 26 (1.1) |
| Smoking history | Never smoked | 1478 (60.7) |
|  | Ever smoked | 959 (39.3) |
|  | Missing data | 0 (0) |
| Partner status | Has partner | 2365 (97.1) |
|  | Does not have partner | 72 (2.9) |
|  | Missing data | 0 (0) |
| Alcohol consumption in pregnancy | No consumption | 2326 (95.4) |
|  |  | <10 |
|  | Missing data |  |
| *Pregnancy and delivery characteristics* |  |  |
| Gestational diabetes | Has gestational diabetes | 105 (4.3) |
|  | Missing data | <10 |
| Hypertensive status in pregnancy | Normotensive | 2182 (89.5) |
|  | Missing data | <10 |
| Gestational age at delivery (weeks + <sup>days</sup>) | ≤36+<sup>6</sup> | 92 (3.8) |
|  | Missing data | <10 |
| Mode of delivery | Vaginal delivery | 1762 (72.3) |
|  | Missing data | <10 |
| *Infant characteristics* |  |  |
| Birthweight centile | <10<sup>th</sup> | 207 (8.5) |
|  | >90<sup>th</sup> | 105 (4.3) |
|  | Missing data | 0 (0) |
| NICU admission | Yes | 110 (4.5) |
|  | Missing data | <10 |
| Significant childhood morbidities* | Yes | 52 (2.1) |
|  | No | 2385 (97.9) |
|  | Missing data | 0 (0) |
| Sex | Male | 1224 (50.2) |
|  | Female | 1213 (49.8) |
|  | Missing data | 0 (0) |
| *Socioeconomic characteristics* |  |  |
| Index of Multiple Deprivation Score (2007) | 1-10 | 1572 (64.51) |
|  | 11-20 | 522 (21.42) |
|  | 21-30 | 215 (8.82) |
|  | >30 | 29 (1.19) |
|  | Missing data | 99 (4.06) |
| Maternal occupation | Managerial/Professional | 1206 (49.49) |
|  | Missing data | 0 (0) |
| School funding type Early years | Academy | 537 (22.04) |
|  | Community | 1140 (46.48) |
|  | Voluntary | 711 (29.18) |
|  | Missing data | 49 (2.01) |
| Key Stage 1 | Academy | 615 (25.24) |
|  | Community | 899 (26.89) |
|  | Voluntary | 542 (22.24) |
|  | Missing data | 381 (15.63) |

For our instrument variable (IV) we selected meta-analysed SNPs that were statistically significant for BMI with a p-value <1x10^-11^ and a linkage disequilibrium (LD) *r*^2^<0.01 in a window of 250kb that were not scaffold variants and were present in the POPS genetic dataset. Included palindromic SNPs with allele frequency ∼0.5 were removed for data harmonisation purposes.

The secondary exposure was maternal pre-pregnancy BMI. Maternal height was measured at the 20-week visit and pre-pregnancy weight was recalled by participants. We treated BMI as both a continuous variable and a categorical variable (<18.5kg/m^2^, ≥18.5-24kg/m^2^, ≥25-29kg/m^2^, ≥30kg/m^2^) for modelling. Participants with missing data were excluded from analysis.

We regressed BMI as a continuous variable on PRS_BMI_ and used the residuals from the model as a proxy for environmentally-driven BMI. We refer to this exposure as ‘non-PRS BMI’ throughout.

We stratified women by their observed BMI and genetic risk for BMI into four risk groups. A PRS_BMI_ z-score >0 was demeaned to represent above average genetic risk within the study population and was thus deemed ‘high risk’ at a cohort level, whereas a PRS_BMI_ z-score <0 represented the opposite. Women with PRS_BMI_ z-score >0 and BMI >25kg/m^2^ were classed as ‘high-risk genotype, high-risk phenotype’ (HG/HP); women with PRS_BMI_ z-score <0 and BMI>25kg/m^2^ were classed as ‘low-risk genotype, high-risk phenotype’ (LG/HP); women with PRS_BMI_ z-score >0 and BMI<25kg/m^2^ were classed as ‘high-risk genotype, low-risk phenotype’ (HG/LP) and those with PRS_BMI_ z-score <0 and BMI<25kg/m^2^ were classed as ‘low-risk genotype, low-risk phenotype’ (LG/LP).

## Statistical methods

Continuous variables were expressed as mean ±SD, while categorical variables were expressed as counts and percentages.

To control for population stratification, we calculated the first 10 genetic principal components (PCs) of ancestry in POPS on typed variants with MAF >0.05, missingness <1%, and that had been pruned for linkage disequilibrium using the “--indep-pairwise” function in Plink (window size of 50, step size of 5, r^2^ threshold of 0.1) (n=72,621). Models were further adjusted for maternal age at study entry. As such, we replicated all applicable adjustments made in the meta-analysed GWAS data^(25)^.

Linear regression was used to define the relationship between PRS_BMI_ and observed BMI. Multivariable logistic regression was used to analyse associations between PRS_BMI_ and educational outcomes, adjusting for identified covariates. Linearity was tested using the Box-Tidwell and likelihood ratio tests. Covariates with limited missingness were imputed using multiple imputation by chained equations. The full imputation methods have been detailed elsewhere^(5)^.

False discovery rate was controlled using the Benjamini-Hochsberg correction at an alpha of 0.05. Q values are presented throughout the results where the dependent variable is an educational outcome. All other modelling is presented with p values.

With input from a paediatrician, a pre-specified dummy covariate was created to indicate if an infant had suffered severe childhood morbidity potentially limiting their engagement with education. Full details of the covariate’s derivation have been previously published^(5)^.

Two-sample MR used the inverse-weighted variance method^(26)^ underpinned by the assumptions that the IV SNPs were associated with the exposure; the SNPs only affected the outcome through the exposure; and that the SNPS were independent of confounding in the exposure-outcome relationship[32341063 (27)]. In view of the first assumption, only SNPs with F statistic >10 were included in the IV. We checked for violation of the final assumption in our sensitivity analysis. Methods for MR sensitivity analysis included (i) MR PRESSO^(28)^, (ii) MR Egger^(29)^, and (iii) the weighted median^(30)^. Heterogeneity was assessed using funnel plots. Pleiotropy was assessed through the MR-Egger intercept p-value and the MR-PRESSO global test.

### Sensitivity analysis

As premature infants are at increased risk of reduced educational attainment compared to those born at full term^(31)^, we pre-specified a sensitivity analysis excluding infants born <37^+0^ weeks’ gestation (n=92) from PRS_BMI_ models. We did not exclude pre-term infants from MR analysis. As prematurity may lie on a causal pathway linking raised BMI to reduced neurocognitive outcomes, their exclusion may attenuate any causal effect.

We further conducted a sensitivity analysis using maternal weight measured at 12-weeks’ gestation to derive maternal early-pregnancy BMI. We regressed early-pregnancy BMI on the PRS_BMI_ and used the residuals as a ‘non-PRS early-pregnancy BMI’ covariate.

All analyses were conducted using R for statistical computing (version 4.6.0; R Core Team, 2026), Python (version 2.7), and PLINK (versions 1.9 and 2.0).

Study approvals were received Cambridgeshire 2 Research Ethics Committee, reference 07/H0308/163; Cambridge Central Research Ethics Committee, reference 18/EE/0036; and the Confidentiality Advisory Board, reference 8/CAG/0024. Participants gave written informed consent antenatally. They were able to opt out of data linkage for educational outcomes.

## Results

Our analytic cohort included 57.8% (2437/4212) of the POPS cohort (Figure 1), and broadly reflected its demographics (Table 1, Supplementary Table 1).

**Figure 1:**
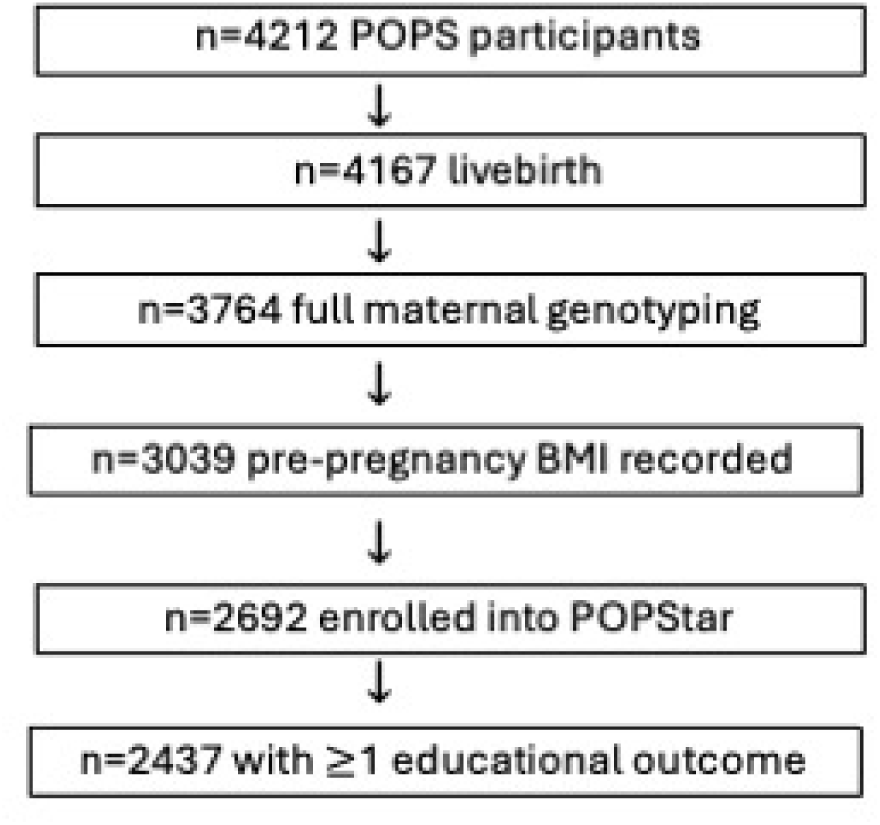
Derivation of analytic cohort.

### (i) Associations between polygenic risk scores, observed BMI, and educational outcomes

The Box-Tidwell and likelihood ratio tests indicated linear relationships between both the maternal BMI and PRS_BMI_ and all educational outcomes.

Each unit increase in maternal pre-pregnancy BMI was associated with ∼6% reduction in the odds of infants attaining GLD (OR 0.94, 95%CI: 0.92-0.97, q<0.001, Table 2). The inverse relationship between observed maternal BMI and infant educational attainment was significant for all further outcomes (Table 2). Results did not materially change when early- instead of pre-pregnancy BMI was used (Supplementary Table 2).

**Table 2:** Odds ratios (95% confidence interval) of infants attaining mid-childhood educational outcomes by i. total observed maternal BMI (body mass index) category, ii. mutually adjusted BMI genetic liability and non-PRS BMI. PRS_BMI_ = polygenic risk score for BMI. Non-PRS BMI = residuals from regression of total BMI on the BMI polygenic risk score. GLD = good level of development. GLD and phonics measured at ages 5-6. All other outcomes measured at age 7. The regression models using total observed BMI were adjusted for maternal factors (maternal age in pregnancy, ethnicity, occupation, partner status, smoking history, first 10 principal components), infant factors (gestational age, sex, birthweight z-score, birth season, serious childhood morbidity), and socioeconomic factors (Index of Multiple Deprivation, school funding type, academic year). PRS_BMI_ and non-PRS BMI were mutually adjusted, with further covariates of maternal factors (maternal age in pregnancy, ethnicity, occupation, partner status, smoking history, alcohol intake in pregnancy, first 10 principal components), infant factors (gestational age, sex, birthweight z-score, birth season, serious childhood morbidity), and socioeconomic factors (Index of Multiple Deprivation, school funding type, academic year).

| Educational outcome | n | BMI | PRS<sub>BMI</sub> | Non-PRS BMI |
|---|---|---|---|---|
| GLD | 2415 | 0.94 (0.92-0.97)<br>q<0.001 | 0.81 (0.72-0.91)<br>q=0.001 | 0.95 (0.93 – 0.98)<br>q=0.001 |
| Phonics | 2382 | 0.93 (0.90-0.97)<br>q<0.001 | 0.77 (0.62 – 0.96)<br>q=0.02 | 0.95 (0.91 – 0.99)<br>q=0.01 |
| Maths | 2055 | 0.95 (0.93-0.98)<br>q<0.001 | 0.76 (0.70 – 0.87)<br>q<0.001 | 0.97 (0.94 – 0.99)<br>q=0.02 |
| Science | 2055 | 0.93 (0.91-0.96)<br>q<0.001 | 0.78 (0.66 – 0.92)<br>q<0.01 | 0.94 (0.91 – 0.97)<br>q=0.001 |
| Reading | 2054 | 0.95 (0.93-0.97)<br>q<0.001 | 0.79 (0.69 – 0.90)<br>q=0.001 | 0.96 (0.93 – 0.99)<br>q=0.01 |
| Writing | 2055 | 0.95 (0.93-0.97)<br>q<0.001 | 0.79 (0.70 – 0.89)<br>q<0.001 | 0.96 (0.94 – 0.99)<br>q<0.01 |

When BMI was categorised, infants of women with BMI<18.5kg/m^2^ were at no greater risk of failing to attain GLD than those of women with BMI 18.5-24kg/m^2^ (Table 3). However, we found a dose-response relationship in infants of women with BMI 25-29kg/m^2^ and BMI ≥30kg/m^2^, with decreasing odds of attaining GLD by increasing BMI category (Table 3). The negative correlation between maternal BMI and likelihood of meeting education standards persisted across all educational outcomes, although did not reach statistical significance for science and maths aged 7 in the BMI 25-29kg/m^2^ category (Table 3).

**Table 3:** Odds ratios (95% confidence interval) of attaining educational outcomes by BMI (body mass index) category. GLD = good level of development. GLD and phonics measured at ages 5-6. All other outcomes measured at age 7. Models adjusted for maternal factors (maternal age in pregnancy, ethnicity, occupation, partner status, smoking history, first 10 principal components), infant factors (gestational age, sex, birthweight z-score, birth season, serious childhood morbidity), and socioeconomic factors (Index of Multiple Deprivation, school funding type, academic year).

| Educational<br>outcome | BMI <18.5<br>n = 96 | BMI 18.5-24.9<br>n = 1548 | BMI 25-29.9<br>n = 556 | BMI≥30<br>n = 237 |
|---|---|---|---|---|
| GLD | 0.87 (0.52-1.53)<br>q=0.63 | reference | 0.67 (0.52-0.87)<br>q=0.01 | 0.52 (0.37-0.75)<br>q=0.01 |
| Phonics | 0.52 (0.23-1.36)<br>q=0.19 | reference | 0.46 (0.29-0.75)<br>q=0.01 | 0.40(0.23-0.73)<br>q=0.01 |
| Maths | 1.28 (0.69-2.57)<br>q=0.49 | reference | 0.79 (0.59-1.07)<br>p=0.17 | 0.57 (0.39-0.83)<br>q=0.01 |
| Science | 2.12 (0.89-6.35)<br>q=0.17 | reference | 0.71 (0.49-1.03)<br>q=0.11 | 0.48 (0.31-0.76)<br>q=0.01 |
| Reading | 1.50 (0.78-3.16)<br>q=0.30 | reference | 0.69 (0.51-0.93)<br>q=0.03 | 0.57 (0.40-0.84)<br>q=0.01 |
| Writing | 1.42 (0.78-2.74)<br>q=0.31 | reference | 0.70 (0.53-0.92)<br>q=0.02 | 0.55 (0.39-0.78)<br>q=0.01 |

Of the variants included in the PRS_BMI_, 91% (1,179,504/1,296,245) were present in POPS genotyping data. PRS_BMI_ explained 19% of the variance in observed BMI (incremental R^2^ = 0.19). Every one standard deviation (SD) increase in PRS_BMI_ was associated with a 2.14 unit increase in observed BMI (95%CI: 1.97-2.32, p<0.001). When non-PRS BMI and PRS_BMI_ were jointly modelled, both were independently associated with decreased odds of meeting expected educational standards aged 5 (non-PRS BMI: OR 0.95, 95%CI: 0.93-0.98, q=0.001; PRS_BMI_: OR 0.81, 95%CI 0.72-0.91, p=0.001, Table 2). This pattern persisted across all educational outcomes (Table 2), and was robust to sensitivity analyses excluding pre-term infants (Supplementary Table 2), and using maternal weight measured in early pregnancy to calculate maternal BMI (Supplementary Table 2).

The infants of women with both high BMI and high PRS_BMI_ (HG/HP) had decreased odds of meeting all educational outcomes compared to all other risk groups (Table 4). Compared to the children of mothers with both low PRS_BMI_ and BMI <25kg/m^2^ (LG/LP), the children of mothers who were discordant in their phenotype/genetic risk (either HG/LP or LG/HP) were also less likely to achieve expected standards, although this only met the threshold for significance in certain educational outcomes (Table 4). Children of mothers with high BMI but low PRS_BMI_ (HG/LP) were less likely to meet standards in maths aged 7, whereas those with low BMI but high PRS_BMI_ (LG/HP) were less likely to meet standards in writing aged 7.

**Table 4:** Odds ratios (95% confidence interval) of attaining educational outcomes by BMI (body mass index) and polygenic risk score category. LG/LP = low-risk genotype, low-risk phenotype. HG/LP = high-risk genotype, low-risk phenotype. LG/HP = low-risk genotype, high-risk phenotype. HG/HP = high-risk genotype, high-risk phenotype. GLD = good level of development. GLD and phonics measured at ages 5-6. All other outcomes measured at age 7. Models adjusted for maternal factors (maternal age in pregnancy, ethnicity, occupation, partner status, smoking history, first 10 principal components), infant factors (gestational age, sex, birthweight z-score, birth season, serious childhood morbidity), and socioeconomic factors (Index of Multiple Deprivation, school funding type, academic year

| Educational outcome | LG/LP<br>n = 962 | HG/LP<br>n = 682 | LG/HP<br>n = 207 | HG/HP<br>n = 586 |
|---|---|---|---|---|
| GLD | reference | 0.89 (0.67-1.18)<br>q=0.46 | 0.70 (0.47-1.07)<br>q=0.14 | 0.57 (0.43-0.75)<br>q<0.001 |
| Phonics | reference | 0.95 (0.54-1.67)<br>q=0.85 | 0.63 (0.30-1.41)<br>q=0.26 | 0.43 (0.25-0.71)<br>q<0.01 |
| Maths | reference | 0.67 (0.49-0.92)<br>q=0.03 | 0.60 (0.39-0.96)<br>q=0.06 | 0.56 (0.40-0.78)<br>q<0.01 |
| Science | reference | 0.77 (0.51-1.15)<br>q=0.23 | 0.61 (0.45-1.12)<br>q=0.14 | 0.49 (0.33-0.74)<br>q<0.01 |
| Reading | reference | 0.80 (0.58-1.10)<br>q=0.21 | 0.63 (0.40-1.01)<br>q=0.09 | 0.55 (0.40-0.76)<br>q=0.001 |
| Writing | reference | 0.76 (0.57-1.01)<br>q=0.10 | 0.61 (0.40-0.93)<br>q=0.04 | 0.54 (0.40-0.73)<br>q<0.001 |

### (ii) Mendelian Randomisation

From the GWAS meta-analysis, n=522 SNPs were included in the IV. The mean F statistic was 85.15. In two-sample MR, there was no difference in the likelihood of children of mothers with high BMI reaching expected educational standards at age 5 or at age 7 across the domains of maths, science, reading and writing (Table 5); all point estimates were negative, however none reached statistical significance. The likelihood of attaining expected standards aged 6 was reduced in children of mothers with high BMI (OR 0.28; 95%CI 0.13-0.62, q=0.01, Table 5). No models showed evidence of heterogeneity or directional pleiotropy (Table 5), with the exception of writing at age 7 where the MR Egger intercept p-value was <0.05. However, the MR PRESSO global test found no outliers and there was no strong evidence of heterogeneity on the funnel plot for this outcome (Table 5).

**Table 5:** Odds ratios (95% confidence interval) of infants attaining educational outcomes per one unit increase in genetically-predicted maternal body mass index on two-sample Mendelian randomisation according to i. inverse variance weighted ii. MR Egger iii. weighted median iv. MR PRESSO methods. Models adjusted for maternal age in pregnancy and the first 10 maternal principal components. GLD = good level of development. GLD and phonics measured at ages 5-6. All other outcomes measured at age 7.

| Educational outcome | n | Inverse variance weighted | MR Egger | Weighted median | MR PRESSO | Funnel plot |
|---|---|---|---|---|---|---|
| GLD | 2417 | 0.84<br>(0.55-1.26)<br>q=0.48 | 0.64<br>(0.19 -2.15)<br>q=0.47<br>Intercept: p=0.64 | 0.60<br>(0.30-1.22)<br>q=0.46 | 1.17<br>(0.79 -1.74)<br>p=0.22<br>No outliers | No evidence of directional pleiotropy |
| Phonics | 2382 | 0.28<br>(0.13-0.62)<br>q=0.01 | 0.12<br>(0.01 -1.22)<br>q=0.09<br>Intercept: p=0.44 | 0.31<br>(0.08-1.21)<br>q=0.09 | 0.29<br>(0.14-0.62)<br>q=0.001<br>No outliers | No evidence of directional pleiotropy |
| Maths | 2056 | 0.84<br>(0.51-1.36)<br>q=0.48 | 1.80<br>(0.42-7.69)<br>q=0.62<br>Intercept: p=0.27 | 1.23<br>(0.52-2.92)<br>q=0.62 | 0.83<br>(0.51-1.32)<br>q=0.62<br>No outliers | No evidence of directional pleiotropy |
| Science | 2056 | 0.70<br>(0.37-1.31)<br>q=0.48 | 0.81<br>(0.13-5.12)<br>q=0.82<br>Intercept: p=0.87 | 0.79<br>(0.28-2.18)<br>q=0.82 | 0.73<br>(0.40-1.35)<br>q=0.82<br>No outliers | No evidence of directional pleiotropy |
| Reading | 2055 | 0.81<br>(0.51-1.30)<br>q=0.48 | 2.48<br>(0.58-10.60)<br>q=0.66<br>Intercept: p=0.11 | 0.90<br>(0.41-1.98)<br>q=0.81 | 0.84<br>(0.52-1.34)<br>q=0.69<br>No outliers | No evidence of directional pleiotropy |
| Writing | 2056 | 0.64<br>(0.42-1.00)<br>q=0.15 | 2.29<br>(0.63-8.36)<br>q=0.20<br>Intercept: p=0.04 | 0.61<br>(0.29-1.25)<br>q=0.20 | 1.50<br>(0.97-2.32)<br>q=0.16<br>No outliers | No evidence of directional pleiotropy |

## Discussion

Our results suggest that maternal pre-pregnancy BMI≥25kg/m^2^ is associated with poorer educational attainment overall, with the strongest and most consistent effects observed in women who also carry a high-risk genotype. Our results suggest that both maternal BMI genetic liability (PRS_BMI_) and the maternal BMI that is not explained by the PRS (the residuals after controlling for genetic BMI) are associated with a reduced likelihood of children achieving expected educational standards in mid-childhood.

To further explore this association we performed MR analysis, with mixed results. Two-sample MR identified a significant inverse relationship in only one of the six mid-childhood educational outcomes reported; the phonics assessment at aged 6. However the point estimates for other educational outcomes also indicated lower attainment, suggesting that the relatively small sample size may have limited the interpretability of this analysis.

Our findings are broadly consistent with previous studies linking increased maternal BMI to adverse neurocognitive development^(2)^,^(3)^,^(4)^ and poorer educational outcomes^(5)^ in children. We further demonstrate that both genetic predisposition to increased BMI, captured by PRS_BMI_, and residual observed BMI unexplained by the PRS are independently associated with decreased educational attainment in mid-childhood. The independent association observed for non-PRS BMI may reflect several potential pathways, including environmental influences on BMI, rare genetic variants, or other sources of BMI variation not represented within the PRS. It is likely that there is, in reality, a complex interplay of biological, behavioural, and environmental mechanisms underlying the relationship. Such mechanisms may include gene–environment interactions^(32)^; indirect genetic effects^(33)^; environmental exposures associated with genetic liability to high BMI^(34)^; and genetic influences not adequately captured by current polygenic scores.

However continuing uncertainty around the causal nature of the association between maternal BMI and offspring educational outcomes limits our ability to make clear recommendations around maternal weight-management interventions specifically to improve long-term offspring outcomes. Several non-MR approaches have previously been used to address residual confounding, including adjusting for paternal BMI^(35)^,^(36)^, paternal negative-control analyses^(15)^,^(37)^ and within-family comparison designs^(38)^. Collectively, these studies have produced conflicting findings, with variable support for a direct intrauterine effect of maternal adiposity on offspring outcomes. Our MR analysis contributes to this ongoing debate but does not fully resolve it. The association between increased maternal BMI and not meeting expected standards in phonic assessment may represent a particularly informative outcome because it captures persistent non-attainment across two serial assessment points (ages six and seven), rather than not meeting standards at a single time point. Consequently, this outcome may identify a subgroup of children with more pronounced educational difficulties, potentially explaining why evidence for a causal effect was strongest for this in two-sample MR.

Notably, in association modelling, the largest effects of BMI on educational attainment were observed among women with both elevated observed BMI and a high genetic liability to high BMI, suggesting that interventions aimed at reducing maternal adiposity could be particularly beneficial within this subgroup. The limited success of previous clinical studies attempting to improve short-term pregnancy outcomes through diet and lifestyle interventions during pregnancy^(39)^,^(40)^ suggests that targeting maternal BMI interventions during the pre-conception phase, for example with GLP1-agonists, could be a more effective and realistic strategy.

Yet whilst manipulation of environmental factors may successfully reduce BMI, it is possible the effects on our outcomes of interest may not be equivalent to the same reduction in BMI achieved through manipulation of genotype (and indicated by MR estimates). Both duration of the exposure and the biological pathways utilised between genetic vs. environmental BMI reduction may not completely overlap, potentially negating an equivalent effect on outcomes ^(41)^,^(42)^.

## Strengths and limitations

We apply a genetic epidemiological approach to examining the relationship between maternal pre-pregnancy BMI and mid-childhood educational attainment. Strengths include the use of a deeply-phenotyped cohort with high-quality genetic data and ∼80% linkage to nationally-standardised mid-childhood education outcomes. A further strength is our ancestry-robust PRS approach, enhancing the generalisability of our results across populations by (i) using a PRS maximising BMI predictive performance across ancestries, (ii) utilising continuous ancestry adjustment when applying the PRS, and (iii) adjusting all PRS and MR regressions for the first 10 PCs.

A key limitation is the use of self-reported weight in the formulation of pre-pregnancy BMI; ∼20% of women in POPS did not have pre-pregnancy weight recorded and were thus excluded from our analysis. However, the baseline characteristics between the whole cohort and our analytic sample were largely similar. A sensitivity analysis using maternal weight measured at 12 weeks’ gestation to calculate early-pregnancy BMI did not materially change our results; and a strong correlation (*r*>90) between recalled and measured pre-pregnancy weight found in the broader literature^(43)^ suggest that both selection and recall bias within our study is limited.

In terms of MR, a key limitation is the potential violation of the MR exchangeability assumption, in which there should be no confounding of the IV-outcome relationship^(44)^. Any assortative mating by BMI could have confounded the pathway between maternal genetic risk and infant outcomes via paternal genotype^(45)^, thus biasing our estimates^(46)^. Another limitation is potential violation of the MR exclusion restriction – whereby the IV should only affect the outcome through the exposure^(44)^ - as residual pleiotropy cannot be ruled out. For example, BMI genetic variants have been associated with impulsivity^(47)^ and reward processing^(48)^. Any analysis that uses these BMI SNPs may thus be related to alternative pathways influencing infant educational attainment through a social effect^(49)^ rather than maternal biological adiposity. Further, these BMI-linked behavioural alleles could affect educational performance via a direct effect in the infant.

Our findings may also be a result of limited statistical power to detect modest effects on educational outcomes. This possibility is particularly relevant for the two-sample MR analyses, where effect estimates were imprecise and confidence intervals were wide. Furthermore, the effects of BMI-associated genetic variants may differ according to demographic and environmental factors such as age and socioeconomic position, even within ancestry groups^(32)^,^(50)^. Such context-dependent genetic effects could reduce both instrument validity and the generalisability of our results.

## Conclusion

We demonstrate an inverse relationship between maternal BMI and mid-childhood educational attainment that did not consistently reach statistical significance in causal analysis, warranting further study in larger cohorts. Resolving these relationships is important as maternal BMI is a potentially modifiable risk factor for childhood educational attainment. A distinct intrauterine causal relationship with observed BMI at the start of pregnancy would suggest potential benefit from pre-conception maternal weight management to improve childhood outcomes.

## Data availability statement

Participant level data is accessed through a Secure Research Service and thus cannot be made publicly available.

