## Supplementary Tables 1-3 for "Maternal genetic risk for high body mass index and reduced infant mid-childhood educational attainment"

| **Characteristic** |  |  |  |
| --- | --- | --- | --- |
| *Maternal characteristics* |  | Analytic cohort (n=2437) | Eligible POPStar participants (n=4164) |
| Age at recruitment (years) |  | 29.80 (5.1) | 29.9 (5.1) |
| Early-pregnancy BMI (kg/m^2^) | Missing data | 24.9 0 (4.6)  0 (0) | 25.1 (4.7)  <10 |
| Self-reported White/European ethnicity | White  Missing data | 2305 (94.6)  26 (1.07) | 3861 (94.3)  25 (2.7) |
| Smoking history | Never smoked  Missing data | 1478 (60.7)  0 (0) | 2475 (59.4)  0 (0) |
| Partner status | Has partner  Missing data | 2365 (97.1)  0 (0) | 4011 (96.4)  0 (0) |
| *Infant characteristics* |  |  |  |
| Gestational age at delivery (weeks) |  | 40.03 (1.7) | 40 (1.7) |
| Birthweight centile | Missing data | 45.03 (25.9)  0 (0) | 45.6 (26) |
| Sex | Female  Missing data | 1213 (49.8)  0 (0) | 2065 (49.6)  <10 |
| Mode of delivery | Vaginal delivery  Missing data | 1762 (72.3)  <10 | 3008 (72.4)  11 (0.3) |
| Birth season | Born in autumn  Missing data | 670 (27.9)  0 (0) | 1123 (27)  0 (0) |
| *Socioeconomic characteristics* |  |  |  |
| Index of Multiple Deprivation Score (2007) | Missing data | 10.23(6.5)  99 (4.1) | 10.3 (6.5)  171 (4.1) |

Supplementary Table 1: Comparison of characteristics between the study analytic sample and the entire POPS cohort. Any numbers <10 are supressed to comply with the Office of National Statistics’ data presentation policy. Data presented as mean(SD) for continuous variables or number(%) for categorical variables.

| Educational outcome | n | Early pregnancy BMI | PRS_BMI_ | Non-PRS early-pregnancy BMI |
| --- | --- | --- | --- | --- |
| GLD | 2415 | 0.94 (0.92-0.96)  q<0.001 | 0.81 (0.72-0.91)  q<0.001 | 0.95 (0.93 – 0.97)  q<0.001 |
| Phonics | 2382 | 0.93 (0.89-0.96)  q<0.001 | 0.77 (0.62 – 0.96)  q=0.02 | 0.95 (0.90 – 0.97)  q=0.001 |
| Maths | 2055 | 0.94 (0.92-0.97)  q<0.001 | 0.77 (0.67 – 0.87)  q<0.001 | 0.96 (0.93 – 0.98)  q=0.001 |
| Science | 2055 | 0.93 (0.90-0.96)  q<0.001 | 0.78 (0.66 – 0.92)  q<0.01 | 0.94 (0.91 – 0.97)  q<0.001 |
| Reading | 2054 | 0.94 (0.92-0.96)  q<0.001 | 0.79 (0.69 – 0.90)  q<0.001 | 0.95 (0.92 – 0.98)  q<0.001 |
| Writing | 2055 | 0.94 (0.92-0.97)  q<0.001 | 0.79 (0.70 – 0.89)  q<0.001 | 0.95 (0.93 – 0.98)  q<0.001 |

Supplementary Table 2: Odds ratios (95% confidence interval) of infants attaining mid-childhood educational outcomes by i. total observed maternal BMI (body mass index) derived from weight measured at 12-weeks’ gestation, ii. mutually adjusted BMI genetic liability and non-PRS early-pregnancy BMI. PRS_BMI_ = polygenic risk score for BMI. Non-PRS early-pregnancy BMI = residuals from regression of total early-pregnancy BMI on the BMI polygenic risk score. GLD = good level of development. GLD and phonics measured at ages 5-6. All other outcomes measured at age 7. The regression models using total observed early-pregnancy BMI were adjusted for maternal factors (maternal age in pregnancy, ethnicity, occupation, partner status, smoking history, first 10 principal components), infant factors (gestational age, sex, birthweight z-score, birth season, serious childhood morbidity), and socioeconomic factors (Index of Multiple Deprivation, school funding type, academic year). PRS_BMI_ and non-PRS early-pregnancy BMI were mutually adjusted, with further covariates of maternal factors (maternal age in pregnancy, ethnicity, occupation, partner status, smoking history, first 10 principal components), infant factors (gestational age, sex, birthweight z-score, birth season, serious childhood morbidity), and socioeconomic factors (Index of Multiple Deprivation, school funding type, academic year).

| Educational outcome | n | PRS_BMI_ | Non-PRS BMI |
| --- | --- | --- | --- |
| GLD | 2324 | 0.80 (0.71 – 0.90)  q=0.001 | 0.96 (0.93 – 0.98)  q<0.01 |
| Phonics | 2277 | 0.77 (0.62 – 0.96)  q=0.02 | 0.95 (0.91 – 0.99)  p=0.02 |
| Maths | 1975 | 0.74 (0.64 – 0.85)  q<0.001 | 0.97 (0.94 – 1.00)  q=0.03 |
| Science | 1975 | 0.76 (0.64 – 0.90)  q<0.01 | 0.94 (0.91 – 0.97)  q=0.001 |
| Reading | 1974 | 0.78 (0.69 – 0.88)  q<0.001 | 0.96 (0.93 – 0.98)  q=0.01 |
| Writing | 1975 | 0.77 (0.68 – 0.87)  q<0.001 | 0.96 (0.94 – 0.99)  q<0.01 |

Supplementary Table 3: Odds ratio (95% confidence interval) of achieving mid-childhood educational outcomes per one unit increase in mutually-adjusted PRS_BMI_ and non-PRS BMI in infants born at full term (≥ 37 weeks’ gestation) (n=2345). PRS_BMI_ = polygenic risk for body mass index. Non-PRS BMI = residuals of total observed BMI regressed on PRS_BMI ._  GLD = good level of development. GLD and phonics measured at ages 5-6. All other outcomes measured at age 7. Models were adjusted for maternal factors (maternal age in pregnancy, ethnicity, occupation, partner status, smoking history, first 10 principal components), infant factors (gestational age, sex, birthweight z-score, birth season, serious childhood morbidity), and socioeconomic factors (Index of Multiple Deprivation, school funding type, academic year).
